# Learning mixed-infection strains from older hosts: a new sampling scheme for malaria epidemiology and population genetics

**DOI:** 10.1101/2025.11.10.25339771

**Authors:** Jiawei Liu, Chaoyu Ding, Nianqiao Ju, Qixin He

## Abstract

**Background:** Malaria is characterized by frequent mixed infections and extremely high strain diversity, against which hosts do not acquire sterile immunity. The genetically divergent strains circulating at the within-host and population levels bring challenges to efficient disease control. Most molecular epidemiological studies investigate diversity patterns of pathogen isolates from children, usually under the age of five, since children suffer from higher rates of mortality and severe symptoms in endemic regions. However, a higher multiplicity of infection (MOI) in children makes it difficult to resolve the underlying strain genetic structure within hosts and populations, bringing obstacles for studying population genetics of malaria field samples.

**Methods:** In this study, we investigate the impact of host age on the inference accuracy of malaria parasite strains and diversity structure, and propose the best age group sampling strategy for molecular epidemiological studies. By integrating an agent-based malaria transmission model with a state-of-the-art computational tool to resolve mixed infections, we compared the SNP-haplotype inference quality from host samples across different age groups and transmission intensities. Inference quality was assessed using four metrics: the recovery of true haplotypes, strain dictionary, shared infections among hosts, and the underlying strain diversity structure from richness estimators. We then validated the inference quality patterns using an empirical dataset from Uganda.

**Findings:** Samples from the old hosts (age>15) show the best inference accuracy compared to young children (age<5) and young children with low levels of mixed infections (age<5, MOI≤3) from multiple aspects. The within-host strain composition is most accurately inferred from old hosts. Regarding strain haplotype dictionary recovery, 91% haplotypes inferred from old hosts are at least with 0.9 identity to the true strains under intermediate transmission intensity, while that percentage drops to 47% in young children. Under high transmission intensity, the percentages of inferred strains with at least 0.8 identity to the true strains are 83% and 31% in old hosts and young children, respectively. Similarly, the sharing network of infections among hosts is most accurately inferred from old hosts. More importantly, strains accurately recovered from old hosts represent frequent strains at the population level. Evaluation of both simulated and empirical datasets indicates that the strain diversity structure inferred from old hosts also yields the best estimates of population-level strain richness.

**Interpretation:** Our results show that malaria parasite isolates from old hosts can provide the most accurate inference of strain haplotypes, shared infections and population-level strain richness. Therefore, a field study with enriched adult sampling in malaria molecular epidemiology helps inform population-level strain composition, infection patterns, and overall diversity, thereby facilitating research on parasite population genetics and improving effective malaria control. Although the traditionally focused young hosts indeed carry more infections, the high levels of within-host mixed infections bring difficulties to strain inference. Instead, old hosts carry fewer strains, but with higher fitness, thus improving the inference accuracy while facilitating practical disease control. Our findings suggest that older hosts may become a new focus for field sampling in future malaria epidemiological research.

**Funding:** This research was partly supported by the Ralph W. and Grace M. Showalter Award and discretionary funds from the Mary J. Elmore New Frontiers Professorship at Purdue University.

**Research in context:** *Evidence before this study:* A search conducted on PubMed on November 7, 2025, for relevant published research which used the terms ((“malaria”[Title/Abstract] OR “falciparum”[Title/Abstract]) AND (“age structure” OR “age group” OR “age pattern” OR “age-specific”) AND (“genetics” OR “var”)), yielded 81 studies between 1978 and the present day, with only 9 published studies in the past 3 years. Most studies focus on age patterns of disease prevalence, the expression of specific genes or alleles (such as those encoding merozoite surface proteins), and the levels of antibodies. Evidence suggests severe symptoms and mortality in younger hosts, that disease prevalence typically peaks among pre-school or school-age children, and that disease prevalence decreases with age. In the meantime, there is still a considerable number of malaria cases among adolescents and adults in regions with low or intermediate transmission intensity. As the burden of malaria shifts towards older hosts in several regions with declining transmission intensity, evidence suggests that older hosts have overall lower levels of mixed infections and carry more distinct parasites. These findings indicate that older hosts can potentially serve as ideal candidates in learning the parasite strain haplotypes, thus facilitating malaria population genetic studies. Nevertheless, only 4 out of the 79 studies characterized malaria parasites at the level of strain haplotypes, and the age patterns of strain genetic structure remain unexplored. A clear understanding of how malaria strain structure relates to host age is still lacking, obscuring the role of older hosts in improving the inference of genetic structure in malaria.

*Added value of this study:* Recent advances in algorithms to resolve strain haplotypes from mixed infections provide a good opportunity to close this research gap to investigate the impact of host age on learning parasite genetics. Our study integrates a state-of-the-art strain haplotype learning algorithm called SNP-Slice and an agent-based model. We evaluate the effect of host age on the inference quality of parasite SNP-haplotypes from various perspectives: within-host strain inference, strain dictionary across host samples, shared infections between host pairs, and population-level strain richness. Our results suggest that sampling older hosts can facilitate more accurate inference of strain haplotypes and genetic structure in populations under the threat of malaria. This suggested sampling scheme implies potential improvements in estimating endemic transmission intensity and disease burden.

*Implications of all the available evidence:* First, our study confirms previous findings from empirical studies that older malaria hosts carry fewer but more distinct parasite strains. Second, our study reveals, for the first time, that sampling older hosts in field studies can provide a more accurate inference of parasite genetic structure. As a result, we propose a new sampling scheme for malaria field studies that emphasizes sampling old hosts. This scheme is in agreement with the viewpoint that the focal host group of malaria control should be shifted from young children to the relatively old hosts. Finally, we propose a new protocol for malaria molecular epidemiology, advocating for surveys and blood sample collection from adult hosts for genetic sequencing, and then applying haplotype reconstruction algorithms, such as SNP-Slice, to resolve strain haplotypes. With this protocol, strains can be learned with reasonably high accuracy, and there is no need to discard multi-clonal samples. To conclude, our findings will facilitate the population genetic studies of malaria epidemiology, as well as the effective drug deployment and disease control of malaria.

## Introduction

One major hindrance to malaria epidemiology is the ubiquity of within-host mixed infections of genetically divergent parasite strains. While most research has focused on resolving mixed infections using computational algorithms, less attention has been given to how sampling design in field studies influences this process. The extremely high polymorphism in antigen-coding gene families, notably in the *var* gene family encoding erythrocyte membrane protein 1,^1,2^ leads to high diversity of strains at population and host levels. Within-host mixed infections have been widely observed and reported in empirical studies, with around 57% of host isolates carrying more than one strain across 153 published studies.^3^ Because malaria parasites are haploid during the blood stage, strain haplotypes from single infections can be readily obtained from sequencing data. In contrast, resolving strain haplotypes in mixed infections remains challenging. Without resolved haplotypes, population genetic research on malaria parasites is limited in its ability to effectively estimate the true disease burden and inform disease control.

Recent advances in computational tools (such as SNP-Slice,^4^ DEploid^5,6^ and PoolHapX^7^) have improved the reconstruction of malaria strain haplotypes from mixed infections. However, their performance declines as population-level infection complexity increases. This makes careful design of sampling schemes in endemic regions essential to reduce infection complexity in field samples, thereby improving the inference quality of parasite strains in malaria epidemiology. An ideal sampling scheme should simultaneously satisfy two criteria: capturing representative parasite strains while maintaining high inference accuracy from computational tools. In practical terms, this means enriching for hosts with lower infection complexity while carrying high-fitness strains, and minimizing inclusion of hosts with highly complex infections of rare strains. Existing evidence indicates that mixed infection rates and parasite fitness vary across host age groups,^8–10^ suggesting that one way to evaluate sampling schemes is to explore the impact of age-biased sampling on learning mixed infections.

Currently, most empirical studies focus on young children due to sampling convenience and clinical concerns,^11–14^ or school-age children, with the peak of prevalence in high-endemic regions.^15,16^ These traditionally sampled young hosts more frequently carry mixed infections of multiple strains, leading to higher levels of complexity and noise in haplotype inference. On the other hand, the relatively understudied adults (here referring to hosts older than the age of 15) usually express lower transmission complexity with smaller MOI,^9,10^ with a potentially higher signal-to-noise ratio in strain inference. Since naturally-acquired immunity to malaria increases with age,^17,18^ strains infecting adults are highly likely to infect children as well, but not vice versa. By comparing host pairwise similarity in antigenic genotypes, older hosts have been found more likely to carry distinct antigens with less mixed infections.^10,19^ Thus, parasite strains infecting adults might be easier to resolve. In contrast, the similar antigenic information carried by young hosts impedes the distinction of separated parasite strains. These findings suggest that adult hosts may satisfy the previously mentioned two optimal criteria of a sampling scheme for studying parasite population genetics in malaria epidemiology. However, the hypothesis that sampling old hosts can facilitate malaria strain inference has yet to be tested and verified, due to the lack of haplotype-level data with age information.

In this study, we test the hypothesis and propose a new field sampling scheme for malaria molecular epidemiology. Instead of the previous focus on clinical and high-prevalence samples, we suggest an adult-focused survey or an age-stratified cross-sectional survey of the local community. We test the validity of our sampling scheme by comparing the inference quality and fitness of SNP-haplotypes from hosts in different age groups. By modifying a developed agent-based model,^20^ we generated simulated malaria transmission datasets under various levels of transmission intensity, recording all infection histories, host ages, and strain haplotypes (see Methods for details). We then performed haplotype reconstruction on host samples from different age groups with the state-of-the-art algorithm SNP-Slice^4^, and evaluated the inference quality of strains from mixed infections and the underlying diversity structure of parasite populations. We found that samples from the hosts older than 15 provide the highest inference accuracy and the closest estimation of population-level transmission intensity, compared to those from young hosts under the age of 5, or the young hosts with low complexity. We then analyzed an empirical dataset from Uganda, where levels of transmission intensity varied among three local populations.^21^ We again found that strain haplotypes inferred from old host samples generated higher estimates of population-level strain richness and successfully reproduced the order of population-wise transmission intensity. Based on these findings, we propose a new sampling scheme for field malaria epidemiology that emphasizes sampling of relatively old hosts over the age of 15. This approach aims to improve inference of circulating strains and transmission intensity from SNP sequencing data, thereby supporting optimized drug deployment and vaccine development for effective malaria control.

## Methods

### Overview

In this modeling study, we employed a modified version of varmodel3,^20^ an agent-based stochastic malaria transmission simulator, alongside SNP-Slice^4^, a recently developed high-performance inference tool for strain haplotype reconstruction. Our aim was to investigate how age biases in host sampling affect the accuracy of inferred strain haplotypes, their fitness profile (measured by population-level strain abundance and strain distinction levels), diversity structure and infection networks. We conducted simulations and compared the outcomes under three different levels of transmission intensity, representative of empirical transmission scenarios ranging from low-prevalence regions (such as South America) to high-prevalence settings (typically found in sub-Saharan Africa). The inference quality was assessed across several metrics: within-host haplotype similarity, reconstruction of strain haplotype dictionary, host pairwise strain sharing network and host pairwise similarity scores. To evaluate the impact of age-biased sampling on estimates of population-level transmission intensity, we computed two widely used strain richness estimators: chao2 and Jackknife2.^22^ These accuracy metrics and diversity estimators represent different aspects of sampling performance: capturing representative parasite strains while maintaining high inference accuracy from computational tools. Finally, we validated our findings using an empirical dataset to determine whether the effects of host age observed in simulations also influence molecular epidemiological inferences in real-world samples. Since the true strains are unknown in the empirical dataset, we focused exclusively on evaluating the impact of host age on estimating the population-level transmission intensity.

### Modeling of malaria transmission and simulation of SNP haplotypes

In the agent-based model, mixed infections of heterogeneous strains within the same host were allowed, with strain-specific immune memory. New strains were dynamically generated by point mutations and recombination events. Parasite *var* repertories, strain frequencies, and infection history were then recorded. We assumed that each SNP site was sampled from one specific *var* gene. Thus, these SNP sites constructed strains showing antigenic polymorphism. We set the number of SNP sites per strain to 45, and assumed that sites were linked in groups of three, providing 15 unlinked SNP regions, which roughly matched the number of chromosomes in *Plasmodium falciparum*. The recombination of these SNP loci followed the same pattern as the corresponding antigen genes in the simulation.

We simulated malaria transmission dynamics with parameters corresponding to low, intermediate and high levels of transmission intensity respectively (scenarios 1-3 hereafter, Table 1). Host population size was kept at 10,000 to represent a local community. Each scenario was replicated 20 times and each simulation was run for 200 years before termination to ensure that the transmission dynamics, antigen and SNP diversity have reached equilibrium. As simulation output, the agent-based model randomly sampled 2,000 hosts with infection status and fully recorded parasite genomic information at the end of the simulation for the downstream analyses. The mean number of infected hosts across 20 replicates for scenarios 1-3 were 313±29·9, 828±33·6 and 1329±26·9 respectively. In the main text, we focus exclusively on infected hosts; therefore, the term population size refers specifically to the number of infected individuals.

**Table 1:** Parameters and epidemiological outcomes for the three simulated scenarios of malaria infection using a modified version of the agent-based model varmodel3.^20^ The parameters not listed below were set to default values from the original version of varmodel3. Parameters for generating different levels of transmission intensity were selected according to Figure 2 (A) in He, Chaillet et al.^23^ The column of transmission potential represents the harmonic mean of daily transmission rate in a seasonal transmission scenario.^20^ The prevalence for each scenario is averaged across 20 simulated populations, with 2,000 as the constant output population size (infected + not infected hosts).

| Scenario | n_genes_initial | n_alleles_per_locus_initial | biting_rate | Transmission potential | Prevalence |
| --- | --- | --- | --- | --- | --- |
| 1: low | 720 | 72 | $2 \times 10^{-5}$ | 0.0183 | $0.157 \pm 0.015$ |
| 2: mid | 2500 | 250 | $5 \times 10^{-5}$ | 0.0458 | $0.414 \pm 0.017$ |
| 3: high | 8100 | 810 | $1 \times 10^{-4}$ | 0.0916 | $0.664 \pm 0.013$ |

### Processing of empirical data

We also inferred SNP haplotypes from SNP calls of malaria isolates collected from three sub-counties in Uganda (Jinja, Kanungu and Tororo) from Chang et al.^21^ Information on host age associated with each isolate was provided by the first author. We applied the same filtering criteria as those used in the original study to remove low-quality samples. Specifically, we excluded host samples with missing data at more than 25% SNP loci and loci with missing information in more than 20% samples. While the original study applied the filter in each population separately, we applied it to the entire combined dataset. The numbers of hosts after filtering were 48, 74, and 462, respectively, in Jinja, Kanungu, and Tororo.

### Age sampling strategies

We first verified the abundance of infections in older hosts. Comparing the number of infected hosts older than 15 years (Old age group) and hosts younger than 5 years (Young age group), we found that the resulting numbers were (in the format of Old & Young): Scenario 1: 124±14·6 & 93·5±12·4; Scenario 2: 273±19·3 & 306±15·2; Scenario 3: 537±18·0 & 367±19·7. These numbers indicate that infections in old hosts are frequent enough to be considered in empirical sampling.

We then generated three age group samples from the infected hosts for studying the effect of host age on strain inference quality: 1) Old age group, 2) Young age group; and 3) Young_sub age group, which was subsampled from Young with MOI no greater than 3. The sample size was kept at 15% of total infected hosts for the Old and Young groups; 128 random sample sets were generated for each scenario. The inclusion of Young_sub was to investigate whether excluding high complexity infections in the Young group can achieve similar performance as that of the Old age group.

The age threshold for the empirical dataset was set differently from the simulation studies because the original study did not apply age-stratified sampling. The distributions of sampled host age were different among the three sub-counties, leading to a lack of enough individuals in certain sub-counties under the previous age threshold for simulations. To ensure each sub-county with enough individuals for each age group, we set top and bottom 30 percentiles of age to define Old (≥ 11) and Young (≤ 5) age groups, resulting in 19, 22 and 145 hosts in the Old age group in Jinja, Kanungu and Tororo respectively, and 11, 21, 160 hosts in the Young age group correspondingly.

### Resolving SNP-haplotypes

For each simulated population with malaria transmission, read counts for major and minor alleles were generated from simulated SNP haplotypes using a Poisson distribution. We then ran SNP-Slice to resolve strain haplotypes using the SS-NegBin method (the negative binomial noise model, designed for high variance in read count), on hosts from the Old, Young, and Young_sub age groups separately to examine the accuracy of strain haplotype inference. For each host sample set, an independent MCMC chain was run for up to 10,000 iterations with a minimum burn-in of 5,000 iterations. After burn-in, the chain may terminate early if the maximum likelihood estimator remains unchanged for 1,000 steps. The concentration parameter α was set to 1, 2 and 3 for Scenarios 1-3, though our previous work showed that inference results were not sensitive to this parameter.^4^ All the other parameters were set to the defaults.

For empirical data with categorical genotype calls instead of read counts, we ran 8 long chains using SS-Cat for Old and Young groups in the combined population of three sub-counties (each continued for a maximum of 20,000 iterations, with 10,000 as burn-in steps, and the chain was stopped after burn-in if the maximum likelihood estimator did not change for 2,000 steps) to get high-quality inferred strain haplotypes from large-scale empirical data.

### Inference quality metrics

1) Within-host inference quality (WHI)

To evaluate the accuracy of resolving mixed infections within each host, we identify the most similar inferred strain to each true strain within the focal host and vice versa. Specifically, we calculate the within-host inference quality (WHI) as follows:

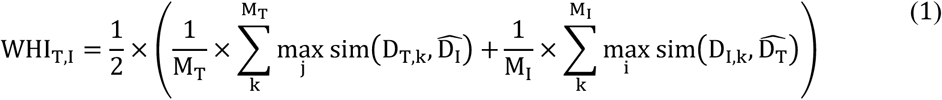

In equation (1), T and I denote true and inferred respectively. D_T_is the true strain haplotype dictionary for the focal host, and M_T_ is the true number of genetically distinct strains (MOI) for this host (D_T_ is a M_T_ × *L* matrix, where *L* is the number of SNP loci). Similarly, D_#_ and M_#_represent the inferred strain haplotype dictionary and MOI for the focal host. For each true strain, we compute its maximum similarity score (sim, defined as the fraction of identical loci shared between two haplotypes) against all inferred strains, then average across the M_!_ true strains. We repeat this procedure in the opposite direction (inferred to true strains). The overall WHI is defined as the mean of these two directional similarity scores.

2) Population level inference quality

We assess the population-level strain inference quality by comparing the similarity of inferred strains to the true strain haplotype dictionary of sampled hosts. For each inferred strain, we find its closest true strain in the subset (with the highest similarity score, defined by the fraction of identical loci) and record similarity score distributions across replicates.

3) Host pairwise strain sharing network

We also evaluate whether the inference correctly recovers strain sharing among hosts. To quantify pairwise sharing, we first construct the true and inferred strain allocation matrix *A*, where rows represent hosts and columns represent strains. Each entry takes a value of 0 or 1, indicating the absence or presence of a given strain in a host. By multiplying the allocation matrix *A* with its transpose, we obtain the host pairwise strain-sharing matrix S. Since the true and inferred dictionary size may differ, it is difficult to compare the inferred number of shared strains with the true numbers. To address this, we binarize S by setting all entries greater than 1 to 1, reflecting only whether hosts share at least one strain. We then evaluate the accuracy of the inferred sharing matrix *S*_1_ against the true sharing matrix *S_T_* by plotting the true positive rate (sensitivity) against the false positive rate (1 – specificity). Points closer to the top-left corner indicate higher accuracy, while points near the diagonal (y = x) correspond to random strain sharing.

4) Host pairwise strain similarity score (PWS)

The host pairwise strain similarity is identical to WHI except that the comparisons are between a pair of hosts instead of true and inferred strains within the same host:

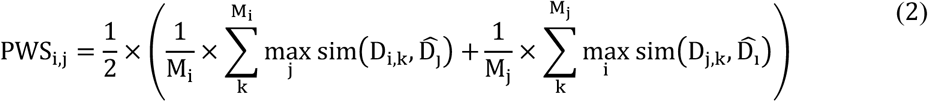

In equation (2), D_’_ is the strain dictionary for host i, and M_’_ is its MOI. For the host pair i and j, we compare each strain in one host to all strains in the other host. For example, in the first half of the terms in the large bracket, host i is the focal host. For each strain in host i, we find the maximum similarity score when comparing it to all strains in host j, and then average across the number of strains in host i. We do the same for the case where host j is the focal host. The pairwise similarity score is then the mean of these two “directional” similarity scores. By obtaining PWS scores across all host pairs using true or inferred strains, we can evaluate the inference accuracy regarding host pairwise strain similarity.

5) Isolate pairwise similarity score (IPS)

When examining the age pattern in signal-to-noise ratios for strain inference, we calculate host pairwise isolate similarity without resolving mixed infections. Specifically, each SNP locus is coded categorically: 0 means the detection of only the major allele and 1 means the detection of only the minor allele, while 0·5 means detection of both alleles. For the loci where two hosts share identical categorical indices (0/0, 1/1, and 0·5/0·5), we assume that they have a similarity of 1. For the loci where one host is homozygous and the other host is heterozygous (0/0·5 and 1/0·5), we define the similarity as 0·5. For the loci where two hosts have opposite indices (0/1), the similarity is naturally 0. The overall host pairwise similarity score is then calculated with equation (3), where L is the number of loci and R is the categorical index. *L_Ri=Rj_* refers to the number of loci where the categorical indices are identical in hosts i and j, and *L*_|*Ri=Rj*|_ = 0.5 represents the number of loci where the allelic status is homozygous in one host and heterozygous in the other host.

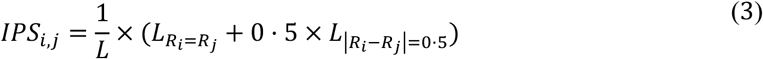

6) Richness estimators

We use two richness estimators, chao2 and Jackknife2, to evaluate how strain structures from host samples can indicate the population-level strain richness. The strain allocation matrix *A* mentioned in part 3 of this section is used as the input to calculate the richness estimators. The estimators are calculated using the functions chao2 and jack2 in the R package fossil.

## Results

### Older hosts carry fewer strains, but with higher fitness

To evaluate the extent to which our agent-based simulations capture empirical transmission dynamics, we quantitatively analyzed simulated datasets to compare infection prevalence and strain composition between Old and Young age groups, including multiplicity of infection (MOI, here defined as the number of genetically distinct strains), population-level abundance of strains, and host pairwise strain similarity. Our results suggested that older hosts typically carry fewer parasite strains, but these strains have overall higher fitness (more abundant in the population with more distinct haplotypes).

MOI distribution of Old was more right-skewed than that of Young, indicating overall lower MOI in Old (Figure 1a). The contrast was more apparent under intermediate or high transmission intensity. In all three scenarios, single infections dominated the Old age group, providing excellent reference information for the inference algorithm to resolve the mixed infections. Conversely, mixed infections dominated the Young age group under intermediate or high transmission intensity, leading to much higher infection complexity.

**Figure 1:**
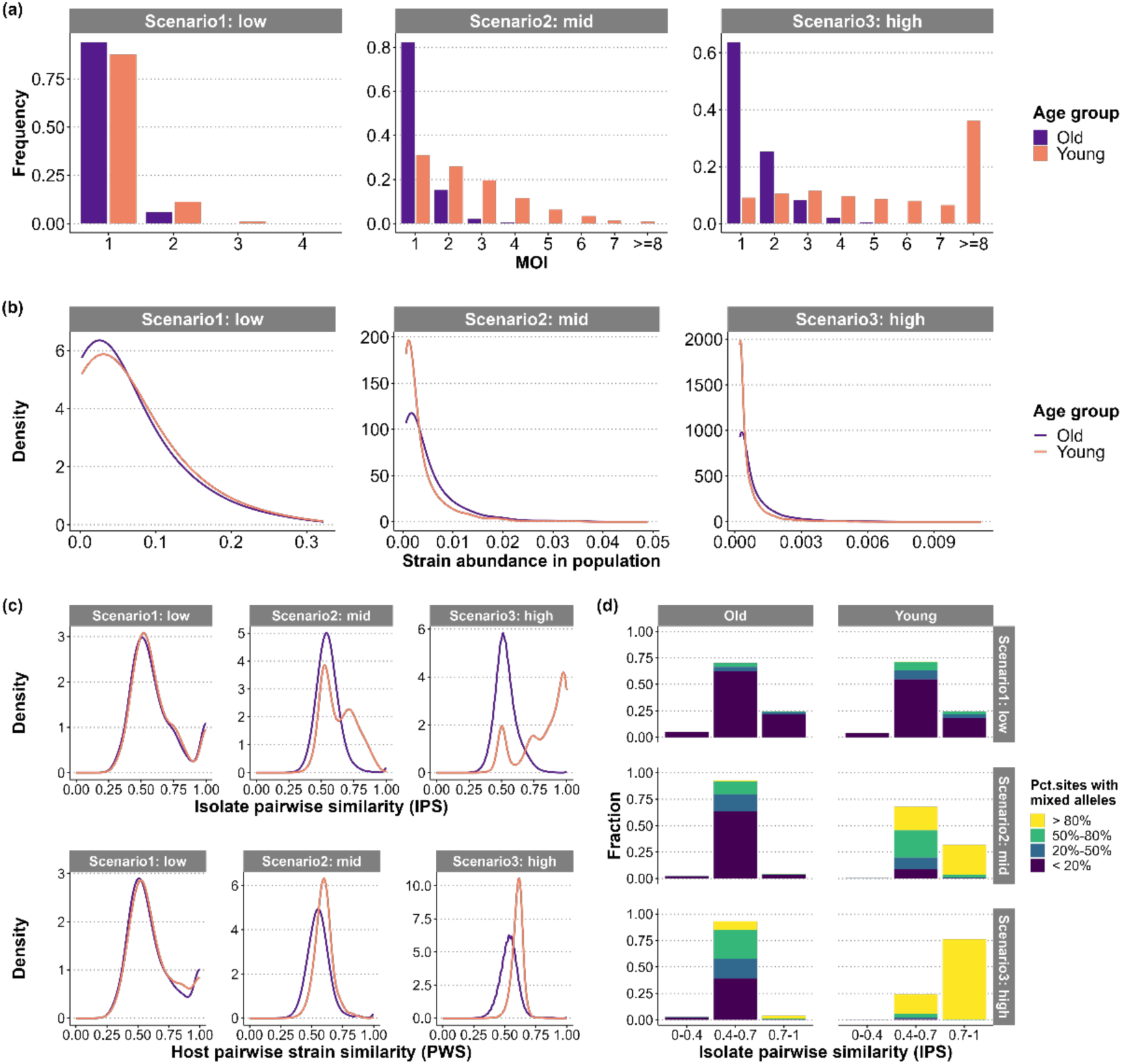
Age structure in infection prevalence and strain composition. 20 populations were simulated under each transmission intensity, and age patterns were assessed by comparing the Old age group (>15 years old) with the Young age group (<5 years old) in the following transmission profiles: (a) The MOI distribution. (b) Distribution of population-level abundance of strains infecting individuals from each age group. (c) Isolate pairwise strain similarity scores (equation (3) in Methods; upper plot) and host pairwise strain similarity scores (equation (2) in Methods; lower plot). (d) Fraction of sites with mixed reads in each host pair, under certain ranges of IPS, based on the upper panel of (c). One locus is regarded as having mixed reads when any one from the host pair is heterozygous (having at least one 0·5 as the categorical index).

Next, we compared the population-level abundance of strains infecting the Old and Young age groups, as a proxy for strain fitness. Most strains infected only a few hosts, shown in the right-skewed strain abundance distribution curves in Figure 1b. Nevertheless, when transmission intensity was moderate or high, Old contained a higher proportion of abundant strains than Young. (Figure 1 (b)). This difference was statistically supported in Scenarios 2 and 3 by one-tailed t-tests (null hypothesis: strain abundance in the Old age group is no higher than that in Young; *p*=0·8146 for Scenario 1; *p*<2·2e-16 for Scenarios 2 and 3).

We then examined pairwise similarities among parasites carried by hosts within each age group and found that older hosts are infected by more distinct malaria parasites. The host pairwise strain similarity (PWS) and isolate pairwise similarity (IPS) scores were significantly lower in Old than in Young with more right-skewed distribution curves, especially under moderate to high transmission intensity (Figure 1 (c), one-tailed t-tests: null hypothesis: scores from Old are no lower than those from Young; PWS: *p*= 0·003 for Scenario 1 and *p*<2·2e-16 for Scenario 2 and 3; IPS: *p*<2·2e-16 for all scenarios). Compared to PWS, IPS was more right-skewed to higher similarity values in Young (top panel in Figure 1 (c)). This pattern arose from the substantially larger fraction of loci with mixed reads between host pairs in Young (Figure 1 (d)). Therefore, the higher IPS in Young reflected extensive mixed infections rather than true genetic relatedness among strains.

In summary, we found that older hosts generally carry fewer but more distinct malaria parasites. These patterns observed in the simulated datasets are consistent with empirical evidence: studies in high-endemic regions have reported lower levels of mixed infections among older hosts.^9,10^ Moreover, older hosts were also reported to carry more distinct polymorphic Duffy-binding-like alpha (DBLα) domains, part of the most important variant surface antigen in malaria pathology.^10^ Therefore, the agent-based simulation successfully captures infection and strain composition patterns. In the following sections, we use simulated datasets to compare the quality of strain haplotype inference across different age groups and provide sampling guidelines based on the results.

### Using older hosts enables higher strain inference quality

To evaluate the effects of age bias in host sampling on strain inference, we compared the inference quality in the Old and Young age groups from several perspectives: within host individuals, between hosts, and at the level of host population. Specifically, we simulated transmission dynamics in one population under each transmission intensity and generated 128 subsampled host datasets per scenario for the following analysis. In addition to Old and Young age groups, we also inspected Young_sub age group by excluding hosts with MOI greater than 3 from Young, to explore the age patterns of inference quality exclusively in hosts with low levels of mixed infections. Basic summary statistics of hosts and strains for the whole population and sampled hosts are listed in Table **2**.

**Table 2:** Summary statistics of the hosts and strains for the simulated populations and host samples used in SNP-haplotype inference. Each scenario is represented by one simulated transmission dynamics and 128 random sets of host samples from the simulated population. #Hosts: the number of infected hosts; #Strains: the number of genetically distinct strains over the entire simulated population; Frac. Hosts: fraction of hosts in each age group relative to the entire population size “#Hosts”; #Sampled hosts: the number of sampled hosts in each age group. #Sampled hosts in Young_sub may vary among sampling replicates and are listed with standard deviations. The columns “#Sampled strains” and “#Inferred strains” represent the numbers of true and inferred strains from the host samples.

| Scenario | #Hosts | #Strains | Age group | Frac. hosts | #Sampled hosts | #Sampled strains | #Inferred strains |
| --- | --- | --- | --- | --- | --- | --- | --- |
| 1: low transmission | 311 | 22 | Old | 0.376 | 47 | 13.1±1.48 | 13.5±1.56 |
|  |  |  | Young | 0.328 | 47 | 11.5±1.13 | 12.6±1.63 |
|  |  |  | Young_sub | 0.328 | 47 | 11.5±1.13 | 12.6±1.63 |
| 2: mid transmission | 846 | 401 | Old | 0.346 | 127 | 109.1±4.63 | 100.5±3.55 |
|  |  |  | Young | 0.349 | 127 | 168.2±7.77 | 120.0±4.13 |
|  |  |  | Young_sub | 0.262 | 95.4±3.87 | 106.7±5.95 | 92.8±5.52 |
| 3: high transmission | 1333 | 2036 | Old | 0.413 | 200 | 263.1±9.51 | 205.9±10.0 |
|  |  |  | Young | 0.267 | 200 | 927.1±27.2 | 184.0±18.0 |
|  |  |  | Young_sub | 0.074 | 55.9±4.52 | 117.5±9.88 | 76.5±8.59 |

At the within-host level, inference quality was highest in the Old age group, reflected both in more accurate MOI estimates (Table S1) and WHI values concentrated near 1 (Figure 2 (a)). The inference quality in Young decreased as transmission intensity increased, suggesting the hindrance of mixed infections to strain haplotype inference. When excluding high MOI hosts in Young, Young_sub showed improved inference quality, but still much worse than Old.

**Figure 2:**
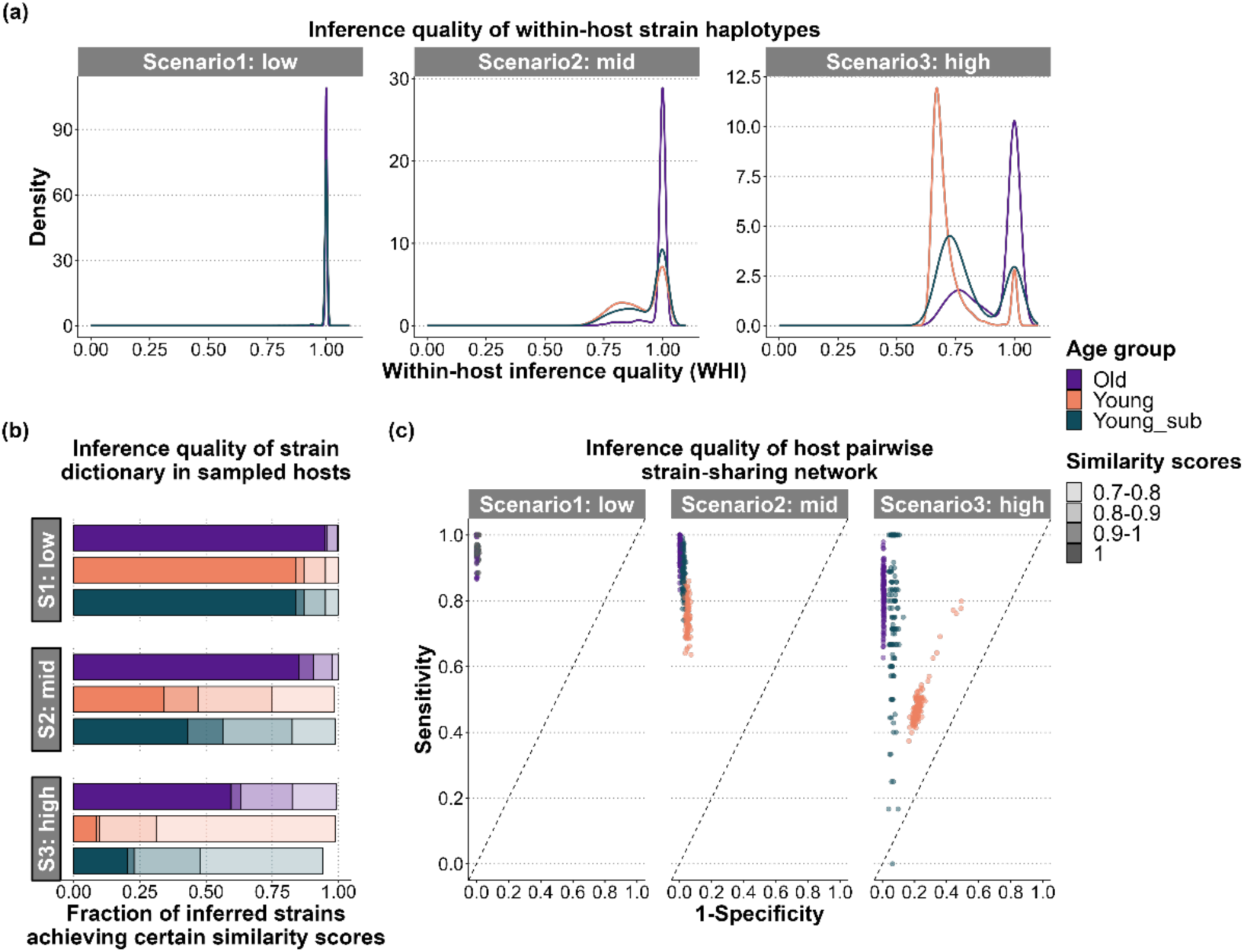
Age patterns in inference quality from different perspectives. For each transmission intensity, one simulated population is picked, and 128 host subsets are sampled from each age group. The identical strains that share all the same SNP loci are merged before any analysis. (a) The within-host inference accuracy of strains, measured by the within-host inference quality (WHI, equation (1)). (b) The inference accuracy of strain dictionary, shown by fractions of inferred strains achieving certain similarity scores, compared to their best matched true strains in the corresponding host samples. (c) The inference accuracy of host pairwise strain-sharing network, shown by plotting the true positive rate (sensitivity) against the false positive rate (1 – specificity).

At the population level, inferred haplotype dictionaries from Old host samples were also closest to the true strain dictionaries, with significantly higher similarity scores (fraction of identical loci) than those from Young (Figure 2 (b), Table S2; p<2·2e-16 across scenarios from one-tailed t-tests with null hypothesis: scores from Old are no higher than those from Young). Although similarity declined with higher transmission intensity, most strains from Old remained highly accurate, with ∼82% ≥0·8 identity and 63% ≥0·9 identity at the highest intensity. In contrast, inference from Young yielded lower fractions of accurately-inferred strains, exacerbated by the smaller inferred dictionary sizes relative to the true dictionaries (Table **2**).

Regarding the between-host strain sharing network, Old again yielded the most accurate inference in terms of binary sharing (Figure 2 (c)) as well as host pairwise similarity scores (PWS; equation (2); Table S3). For the binary host pairwise strain sharing network, we evaluated the accuracy by comparing true positive rate (sensitivity) against false positive rate (1-specificity), where positive stands for sharing strains. While Old produced an accurate network structure across random samples, Young resembled a random sharing network, and Young_sub had a wide variation in inference quality across different samples (Figure 2 (c)).

In summary, across within-host, population, and between-host levels, strain inference was consistently most accurate when sampling Old hosts. The fewer but more distinct malaria parasites carried by old hosts provide better reference strain information and facilitate more accurate inference.

### Inference from older hosts improves recovery of population-level strains

In this section, we went beyond the scope of host samples and assessed whether inferred strains from host samples recover strains, especially frequent strains, at the population-level in different age groups. Because it is not feasible to sample all the circulating strains in a field survey, finding the age group with the best recovery of population-level epidemiological features is essential for increasing the efficiency of malaria field studies.

Under intermediate and high transmission intensities, host samples from the Old age group contained fewer strains than those from Young, reflecting their lower levels of mixed infection (translucent bars in Figure 3 (a), Table S4). Nevertheless, Old achieved the highest recovery of population-level strains from inference (solid bars in Figure 3 (a), Table S5), because of the higher inference accuracy, as we revealed in the previous two sections. The comparable recovery in Young and Young_sub (solid bars in Figure 3 (a)) indicated the restricted role of low MOI young hosts in improving the recovery of population-level strains.

**Figure 3:**
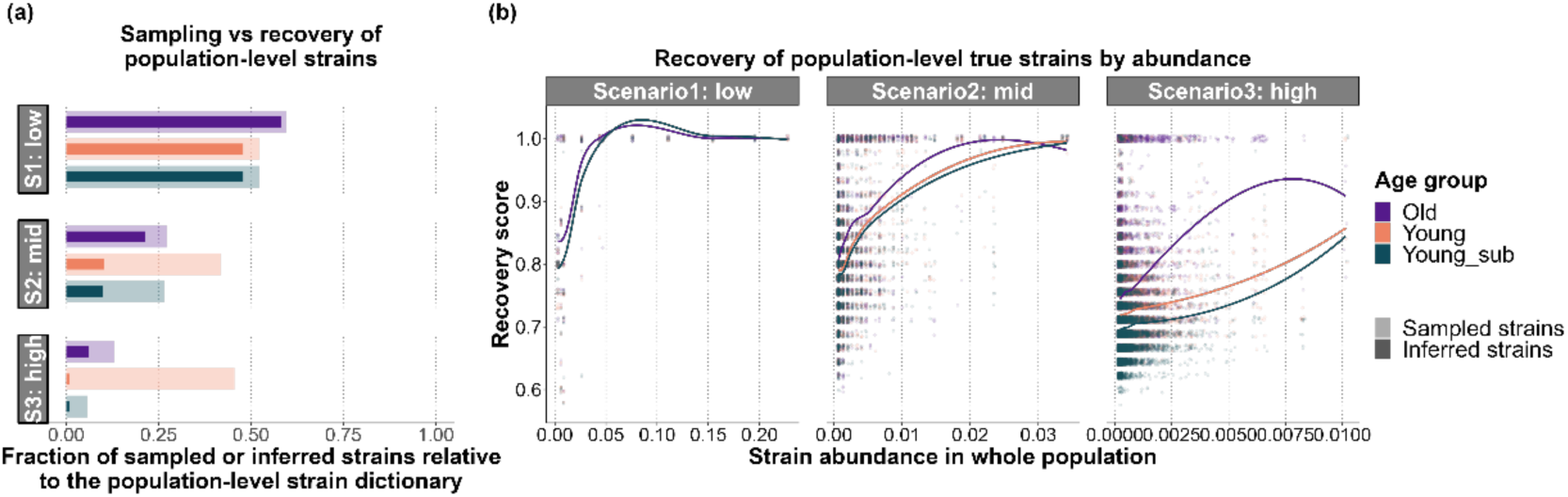
Age patterns in recovery of population-level true strains. Figure 2(a) Sampling and inference recovery of population-level strains in the host samples. The plot shows fractions of strains over the entire population that get included in the randomly selected host subsets (translucent bars) or get completely recovered from SNP-Slice inference of these host samples (solid bars). (b) Relationship between strain abundance and recovery score (similarity of true strains to their best-matched inferred strains). For better visualization, only 5% of the points in each age group are shown in each scenario. The smooth curves are generated by the method loess in the R function geom_smooth with all points.

Next, we evaluated the relationship between strain abundance and recovery score in different age groups. The recovery score increases with strain abundance in all three age group samples. Yet, Old still has higher strain recovery scores than Young and Young_sub across the entire abundance gradient (Figure 3 (b), Figure S1, Table S6), especially at intermediate and high abundance.

The comparisons in this section indicate that the strains inferred from the Old age group are the most representative of population-level high-fitness strains. Although old hosts do not carry as many strains as young hosts, the higher inference quality strengthens their roles in learning the population-level strains.

### Inference from older hosts improves estimation of population-level strain richness

In this section, we explored the age patterns in estimating the population-level strain diversity structure from host samples. We calculated two richness estimators: chao2 and Jackknife2, as applied in a previous study,^22^ to evaluate how strain allocation within sampled hosts can reflect the population-level strain richness (the total number of circulating strains).

We first compared which richness estimators can recover population-level strain richness from true strain dictionary and allocation information (i.e., without inference errors). We found that chao2 statistics in general recovered the correct population richness from all three age groups (Figure 4 (a)). Performance from Young was the most consistent with small variation under medium to high transmission scenarios, with slight overestimation. Old and Young_sub, on the contrary, slightly underestimated strain richness. The other estimator, Jackknife2, generally showed larger underestimates of strain richness except in Young. We therefore focus our discussion on the chao2 estimates for the following comparisons of inferred strain structures.

**Figure 4:**
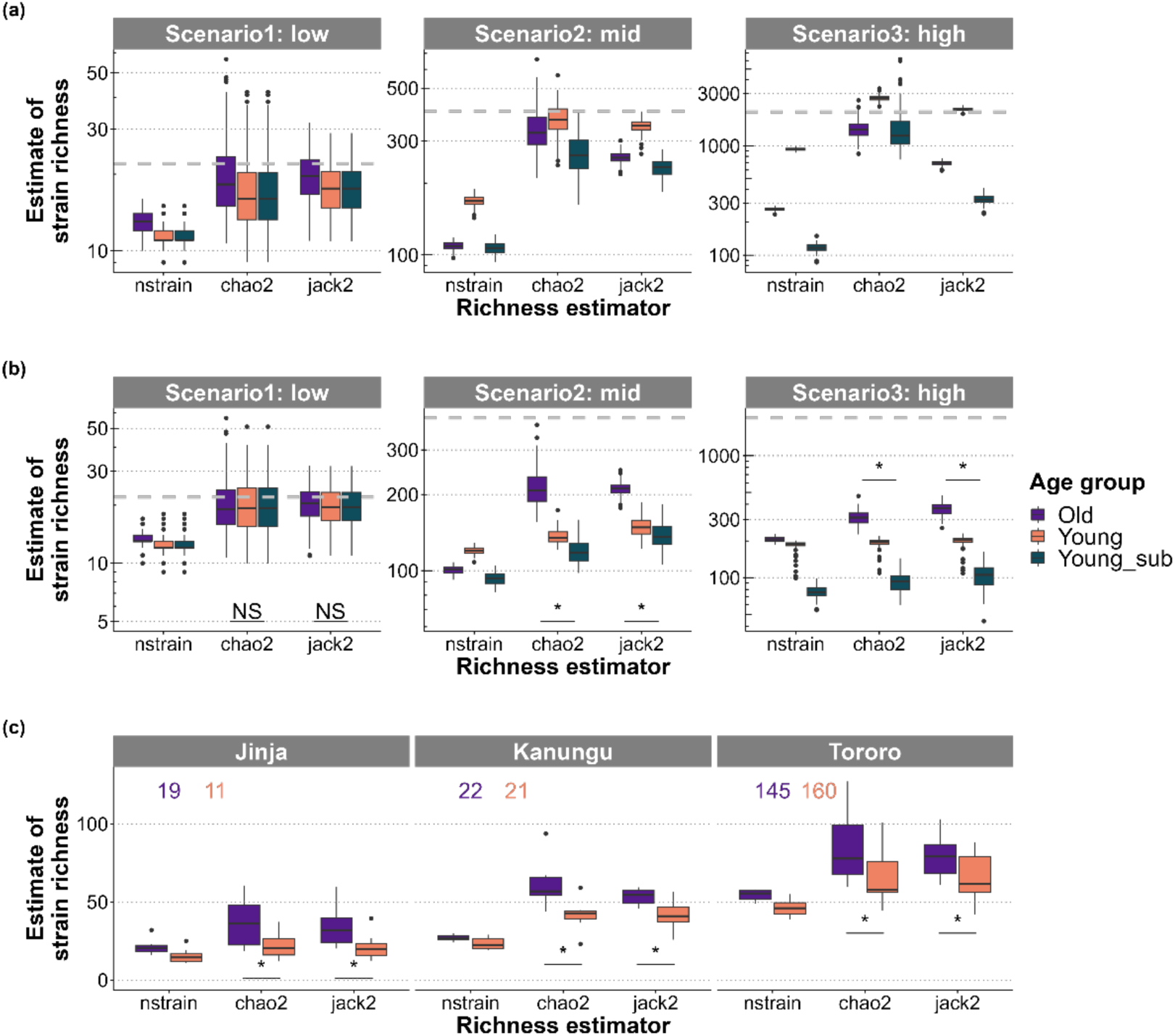
Age patterns in strain richness estimation. (a) Number of strains and richness estimators from the true strain allocation profiles in sampled hosts from simulated populations. The host samples are identical to those in Figure 2 and Figure 3. The grey dashed lines represent the number of strains over the entire population. (b) Number of strains and richness estimators indicated by the inference results, corresponding to the sampled host groups in (a). Asterisks show significantly higher strain richness values estimated from the Old age group compared to those from Young, and “NS” means values from Old are not significantly higher. (c) Number of inferred strains and strain richness estimates of three sub-counties in Uganda. The colored numbers at the top of plots tell the number of hosts in each age group at each location.

Interestingly, based on the inferred results, Old generated the closest estimations to the population-level strain richness (Figure 4 (b)), significantly higher than those from Young under intermediate and high transmission intensity (one-tailed t-test with null hypothesis: strain richness estimator from Old is no higher than that from Young; *p*<2·2e-16 for both estimators under scenarios 2-3; for scenario 1, *p*=0·4431 and 0·07295 respectively). Compared to Figure 4 (a), the estimators derived from inferred strain allocation were lower than those based on true strain allocation, partly due to smaller inferred dictionary sizes (**Table 2**). This contraction was particularly pronounced in the Young group, leading to a deterioration in strain richness estimation. While estimators from both Old and Young qualitatively reproduced the expected order of transmission intensity, Old provided substantially more accurate approximations of true strain richness.

We further tested whether the same patterns held in empirical malaria transmission data. Categorical SNP haplotype information of malaria isolates from three locations in Uganda, together with the host age information, were obtained from Chang et al.^20^ and personal communications. Following the procedure described in Methods, we selected Old and Young hosts based on age percentiles instead of absolute age cutoffs. Inferred strain allocation from Old again provided significantly higher estimates of strain richness (Figure 4 (c), one-tailed t-test with null hypothesis: estimators from Old were no greater than those from Young; *p*<0·05 for all scenarios). Moreover, the inferred results reproduced the expected order of transmission intensity Jinja < Kanungu < Tororo during the time period 2012-2013, when the data was collected.^24^

By analyzing both simulated and empirical malaria infection data, we demonstrated that sampling older hosts enables a more accurate approximation of population-level strain richness and diversity structure. In epidemiological field studies, where true strain allocation is rarely available, inference on old hosts can therefore yield more reliable indicators of the transmission intensity.

## Discussion

In malaria field studies, the widespread mixed infections impede the epidemiological and population genetic research. With restricted parasite strain haplotype and allocation information available, it is necessary to carefully design sampling schemes in endemic regions to efficiently resolve strain structures for the downstream analyses. In this work, we propose a new field sampling scheme that prioritizes sampling from old hosts (adults or post-school-age adolescents) to accurately learn parasite strains from mixed infections. Old hosts carry more distinct high-fitness parasite strains at lower levels of mixed infections. From inference quality comparisons, we demonstrated that malaria genetic epidemiological profiles are the most accurately inferred or estimated from old host samples, ranging from within-host strain haplotypes, strain dictionary, shared infections between hosts to population-level strain richness. Therefore, this new adult-biased sampling scheme simultaneously satisfies the two criteria we mentioned earlier in Introduction: capturing representative parasite strains while maintaining high inference accuracy.

Our suggested sampling scheme is opposite to the frequently applied children-biased sampling scheme in previous malaria epidemiological research. Due to the higher sample abundance under higher symptom and hospitalization rates in children,^25–28^ the traditional sampling scheme focusing on young hosts is useful in quickly assessing the regional disease burden, but the high levels of mixed infections impede the population genetic and epidemiological studies to explain the observed transmission heterogeneity. Practically, the sequencing results might be strikingly similar among isolates from young hosts when strain haplotypes are not resolved. Meanwhile, mixed infections in young hosts bring difficulties to the accurate strain inference. On the other side, isolates from old hosts are better suited for field sampling for strain haplotype reconstruction because of the higher signal-to-noise ratio. In reality, as control measures have overall lowered transmission intensity, malaria burden is tending to shift to older hosts with the decline of transmission intensity,^26,27,29^ further raising the practical importance of our sampling scheme.

This sampling scheme is designed after the epidemiological characteristics and strain evolution patterns of malaria. In young children, the age group that is the most related to clinical malaria, as well as school-age children with the peak prevalence in high endemic regions, the fitness cost of host death and the fitness benefits of faster transmission balance each other, intensifying the total infection with higher levels of mixed infections.^30^ In adult hosts, naturally acquired immunity to malaria increases with age,^17,18^ reducing the mean MOI. Meanwhile, higher strain-specific and cross immunity in adults select for parasites with higher fitness (e.g., higher virulence or higher antigenic diversity).^8^ Children may suffer additional mortality from virulent strains with the presence of asymptomatic adults. Therefore, strains inferred from adults can make significant contributions to real-time disease control not only because of the accurate inference, but also because of their potential high virulence and fitness.

Combining our sampling scheme with the state-of-the-art haplotype-reconstruction algorithm SNP-Slice, we form a complete protocol for future malaria population genetic research. First, we suggest age-stratified sampling with an emphasis on older infected hosts. Genetic materials and data can be obtained by sequencing, and inference by SNP-Slice can generate strain genetic and diversity structures. Since SNP-Slice stays robust for low-concentration or degraded parasite DNA,^4^ our methodology is appropriate for analyzing samples collected from adults or asymptomatic hosts. By combining a specifically designed sampling scheme with an accurate inference algorithm, this protocol aims to maximize the use of the collected samples and minimize the impact of mixed infections, addressing concerns raised by earlier studies that excluded multi-clonal infection samples.^31–38^ Moreover, this protocol enables direct learning of strain structures for further population genetic analyses, instead of calculating secondary quantities or performing principal component analysis.^39–42^ Other than focusing on specific polymorphic antigen genes,^43–45^ or on selected loci relevant to antigens and drug resistance^46–48^ and well-referenced short sequences,^49–52^ accurately learning haplotypes with high population-level fitness is practically important for malaria burden evaluation and vaccine design.

Here we also discuss the limitations of our current protocol. Given fixed research budget, one concern lies in the sufficiency of infected adults due to the generally low prevalence. To address this concern, we balance the tradeoff between sampling convenience and strain inference accuracy. Compared to the traditional clinical-case-biased sampling scheme, reaching the same sample size of old infected hosts may require more sampling efforts, but microscopy can be performed on the blood samples to quickly detect the presence of infections, which has long been a cost-friendly and efficient way for malaria diagnosis.^53^ On the other hand, for samples collected from young children, microscopy is not capable of distinguishing the level of mixed infections to take the subset of individuals with relatively low infection complexity (e.g. microscopy fails to distinguish isolates with MOI=3 and MOI=10). These diagnosed samples with mixed infections would downgrade the accuracy of strain inference other than being discarded, decreasing their contributions to parasite genetic analyses. Therefore, it is reasonable to estimate that the cost of extra sample collection and microscopy on old hosts is affordable, compared to the potential waste on sequencing highly mixed-infected samples from young hosts with little yield of accurate parasite strains. Thus, the new sampling scheme suggested in our study is empirically both effective and efficient for malaria population genetic studies.

Our reported inference quality and population-level strain recovery are still suboptimal under high transmission intensity, a limitation that is inevitable given the non-identifiability of strain haplotypes under high recombination rates. Potential improvements could be achieved by increasing sample sizes or restricting the analysis to more frequent haplotypes. In our simulations, we deliberately incorporated high levels of antigen polymorphism to better illustrate age patterns in inference quality and strain recovery. Inference based on genomic regions with lower complexity would likely face fewer challenges.

Broadly speaking, the sampling scheme and protocol we suggest in this article can also be applied to other pathogens with mixed infections. Infection by genetically distinct strains of the same pathogen has been widely observed in a large variety of pathogen taxa, including bacteria, viruses, protozoa and fungi, where more than 50 human disease pathogens are involved.^55^ Our sampling scheme for malaria should improve efficiency of learning strains in mixed infection for those diseases as well, as long as the natural acquired immunity increases with age. Even if the age pattern in immunity is different in some other diseases, our methodology of comparing strain inference quality from different host groups is applicable for improvement in sampling scheme design by locating the optimal host group. Thus, one future extension will be to apply or adjust our protocol to find out age patterns in strain inference in other infectious diseases with mixed infections, to facilitate the control of these diseases.

In conclusion, we demonstrate that strain haplotype information relevant to molecular epidemiology of malaria can be better learned from older hosts. Meanwhile, strains carried by these old hosts are also potentially the more abundant and virulent ones in the entire population with higher fitness. Therefore, we suggest a new epidemiological sampling scheme to focus on or at least include old hosts. With the integration of strain haplotype resolving algorithm a complete protocol has been constructed to facilitate the analyses of population genetics and host-pathogen landscapes, to bring insights into the real-time disease control of malaria.

## Contributors

JL: conceptualization, data analysis, software, visualization and writing. CD: data cleaning, software, visualization and writing. QH: conceptualization, methodology, software, visualization and writing. NJ: conceptualization, methodology, software, visualization and writing. ·

## Data sharing

All data and code used in this study are publicly available at GitHub: https://github.itap.purdue.edu/HeLab/malaria_age_infer, or can be accessed through the sources cited in the manuscript. Data processing methodologies are available upon request.

## Acknowledgements

We thank Hsiao-Han Chang for providing the age information of hosts in the empirical data from Uganda. We thank the High-Performance Computing support from the Rosen Center for Advanced Computing at Purdue University and the Purdue Institute of Inflammation, Immunology, and Infectious Disease.

