## Supplementary Information for "Learning mixed-infection strains from older hosts: a new sampling scheme for malaria epidemiology and population genetics"

### MOI inference accuracy

Here we list the inference accuracy of MOI in Table S1. For true MOI, the sampled host subsets from one simulated population generally show the representative features in Figure 1(a) in the main text, where 20 populations are assessed under each scenario. The hosts sampled from the Old age group have an overall smaller MOI compared to those from the Young age group, and MOI increases with transmission intensity within each age group. The inference of MOI is overall the most accurate in the hosts sampled from Old, with the smallest root mean squared error (RMSE). More than 80% hosts still have inferred MOI within ±1 from true MOI, except those from Young under high transmission intensity.

**Table S1:** Age patterns in MOI inference, shown by mean and standard deviation of true MOI (M) and inferred MOI ($\widehat{M}$), percentage of individuals with exact MOI recovery, percentage of individuals with MOI error ≤ 1, bias, RMSE and relative error (RSS/TSS). Percentages are calculated after combining inference results of all 128 host samples in each age group under each scenario.

| Scenario | Age group | $\overline{M_i} \pm sd$ | $\overline{\widehat{M}_i} \pm sd$ | % $M_i =$ | % $\lvert\widehat{M}_i - M_i\rvert \leq 1$ | Bias | RMSE | RE |
|---|---|---|---|---|---|---|---|---|
| 1: low | Old | 1·11±0·31 | 1·09±0·37 | 98·9 | 99·1 | 0·019 | 0·103 | 0·577 |
| | Young | 1·13±0·45 | 1·19±0·58 | 97·5 | 97·8 | 0·047 | 0·302 | 0·554 |
| | Young_sub | 1·13±0·45 | 1·19±0·58 | 97·5 | 97·8 | 0·047 | 0·302 | 0·554 |
| 2: mid | Old | 1·28±0·50 | 1·25±0·60 | 96·3 | 98·6 | 0·028 | 0·283 | 0·293 |
| | Young | 2·62±1·50 | 3·06±1·97 | 69·1 | 80·7 | 0·403 | 1·060 | 0·409 |
| | Young_sub | 1·96±0·80 | 2·17±1·28 | 80·9 | 87·7 | 0·320 | 0·810 | 1·050 |
| 3: high | Old | 1·47±0·81 | 1·69±1·13 | 88·2 | 95·8 | 0·162 | 0·531 | 0·420 |
| | Young | 7·19±4·75 | 6·52±3·03 | 28·6 | 51·5 | -0·552 | 2·950 | 0·399 |
| | Young_sub | 2·22±0·85 | 2·69±1·34 | 66·4 | 87·0 | 0·478 | 0·892 | 1·110 |

### Details of comparing strain inference quality in different age groups

Here we provide more details on the comparison of strain inference quality in different age groups. First, we show a summary table of similarity between each inferred strain and its closest true strain in the sampled hosts in Table S2, which is used to plot Figure 2(b) in the main text. For each age group in each scenario, the percentages of inferred strains achieving certain similarity cutoff values are calculated across 128 sets of host samples. Hereafter, whenever we calculate such percentages of strains reaching certain similarity or recovery score, we do that in the same way by combining the results of all 128 host samples.

**Table S2:** The average similarity scores of inferred strains to their closest true strains in the sampled hosts, and percentages of inferred strains reaching certain similarity scores. Percentages are calculated after combining the inference results of all 128 host samples in each age group under each scenario.

| Scenario | Age group | Average | 1 | ≥ 0·9 | ≥ 0·8 | ≥ 0·7 |
|---|---|---|---|---|---|---|
| 1: low | Old | 0·993 | 95·0% | 95·8% | 99·5% | 99·9% |
| | Young | 0·974 | 83·9% | 87·0% | 95·0% | 99·9% |
| | Young_sub | 0·974 | 83·9% | 87·0% | 95·0% | 99·9% |
| 2: mid | Old | 0·980 | 85·1% | 90·6% | 97·6% | 99·9% |
| | Young | 0·884 | 34·1% | 47·0% | 75·0% | 98·3% |
| | Young_sub | 0·907 | 43·0% | 56·4% | 82·5% | 98·9% |
| 3: high | Old | 0·922 | 59·4% | 63·1% | 82·5% | 99·2% |
| | Young | 0·787 | 8·45% | 9·7% | 31·1% | 98·8% |
| | Young_sub | 0·820 | 20·4% | 22·9% | 47·9% | 94·0% |

We also calculate the host pairwise strain similarity scores between all host pairs using equation (3) in Methods, and compare values obtained using true and inferred strains, as an additional measure of between-host level inference accuracy. Table S3 demonstrates that host pairwise strain similarity is most accurately inferred from host samples in Old, suggesting that sampling from relatively old hosts can facilitate estimation of between-host transmission complexity and shed light on host-relatedness in infection.

**Table S3:** Age patterns in inference of host pairwise strain similarity scores: mean and standard deviation of true host pairwise similarity (PWS) and inferred host pairwise similarity ($PWS_I$), bias, RMSE and relative error (RSS/TSS), across 128 host samples in each age group under each scenario.

| Scenario | Age_group | $\overline{\mathbf{PWS}} \pm \mathbf{sd}$ | $\overline{\mathbf{PWS_I}} \pm \mathbf{sd}$ | Bias | RMSE | RE |
|---|---|---|---|---|---|---|
| 1: low | Old | 0·577±0·209 | 0·577±0·209 | 0·000126 | 0·00570 | 0·000742 |
| | Young | 0·591±0·196 | 0·592±0·196 | 0·000425 | 0·0107 | 0·00297 |
| | Young_sub | 0·591±0·196 | 0·592±0·196 | 0·000425 | 0·0107 | 0·00297 |
| 2: mid | Old | 0·559±0·0871 | 0·559±0·0871 | 0·000251 | 0·0159 | 0·0334 |
| | Young | 0·608±0·0745 | 0·611±0·0753 | 0·00281 | 0·0329 | 0·195 |
| | Young_sub | 0·590±0·0776 | 0·593±0·0788 | 0·00296 | 0·0293 | 0·142 |
| 3: high | Old | 0·526±0·0698 | 0·529±0·0707 | 0·00256 | 0·0275 | 0·156 |
| | Young | 0·602±0·0422 | 0·608±0·0500 | 0·00636 | 0·0404 | 0·915 |
| | Young_sub | 0·550±0·0539 | 0·559±0·0633 | 0·00965 | 0·0434 | 0·649 |

### Population-level true strain recovery by sampling or inference from sampled hosts

Here, we provide more details on the recovery of true strains of the entire population from the sampled hosts. Table S4 provides the similarity profile between true strains from the entire population and sampled hosts. The hosts sampled from the Young age group carry the largest subset of parasite strains, as more than 40% of population-level strains are included in the sampled hosts, which account for 15% of the total infected hosts. Moreover, the sampled strains can represent 70%-80% of strains over the entire population, if we take 0·8 as the similarity cutoff value. Conversely, such a percentage drops to around 30%-60% if we sample old hosts under intermediate and high transmission intensity. If we discard young host samples with high levels of mixed infections, the fraction of sampled strains drastically reduces, especially under the highest transmission intensity. The majority of strains infecting young hosts can be excluded if we discard samples with MOI>3.

**Table S4:** The average similarity scores of true strains from the entire population to their closest strains included in the sampled hosts, and percentages of population-level strains reaching certain cutoff similarity levels when compared to the closest sampled strains. Percentages are calculated after combining comparisons over all 128 host samples in each age group under each scenario.

| Scenario | Age group | Average | Similarity =1 | ≥ 0·9 | ≥ 0·8 | ≥ 0·7 |
|---|---|---|---|---|---|---|
| 1: low | Old | 0·907 | 59·7% | 59·7% | 84·2% | 93·4% |
| | Young | 0·886 | 52·2% | 52·2% | 75·7% | 91·4% |
| | Young_sub | 0·886 | 52·2% | 52·2% | 75·7% | 91·4% |
| 2: mid | Old | 0·848 | 27·2% | 30·9% | 63·6% | 94·4% |
| | Young | 0·891 | 41·9% | 46·4% | 79·5% | 97·1% |
| | Young_sub | 0·851 | 26·6% | 30·9% | 65·6% | 94·5% |
| 3: high | Old | 0·783 | 12·9% | 14·2% | 31·2% | 86·7% |
| | Young | 0·886 | 45·5% | 47·5% | 70·5% | 99·6% |
| | Young_sub | 0·737 | 5·77% | 6·48% | 15·8% | 62·6% |

However, the sampled strains may not always be effectively inferred, especially in young hosts with high levels of mixed infections. The percentages of true strains recovered to certain cutoff values by inference are summarized in Table S5. The recovery scores of true strains (similarity of true strains to their closest counterpart in inferred strains) are generally higher if strains are inferred from hosts in the Old age group, indicating the recovery of an overall larger proportion of population-level strains. By running the inference algorithm on host samples from Old, at least 60% of true strains are recovered to the similarity of no less than 0·8 when the transmission intensity is not too high, and about 80% of true strains are recovered to at least 0·7 identical regardless of transmission intensity. We verify the larger recovery scores from Old with one-tailed t-tests, where the null hypothesis is that recovery scores from Old are no higher than those from Young. Results reject the null hypothesis with $p$=2·2e-14 for scenario 1 and $p$<2·2e-16 for scenarios 2-3. The recovery scores in Young_sub are even lower than those in Young, mainly due to the lower coverage of true strains after excluding hosts with large MOI. The percentages in Table S5 look much lower than those in Table S2, because the size of true strain dictionary covering the whole population is always much larger than inferred dictionaries covering only part of hosts, so that we cannot expect the recovery of every true strain from a relatively small fraction of hosts that get sampled.

**Table S5:** The average recovery scores of true strains over the entire population to their closest inferred strains from the sampled hosts, and percentages of population-level strains reaching certain cutoff levels of recovery scores when compared to the inferred strains. Percentages are calculated after combining inference results of all 128 host samples in each age group under each scenario.

| Scenario | Age group | Average | 1 | ≥ 0·9 | ≥ 0·8 | ≥ 0·7 |
|---|---|---|---|---|---|---|
| 1: low | Old | 0·905 | 58·2% | 58·4% | 84·1% | 93·3% |
| | Young | 0·879 | 47·9% | 49·3% | 75·0% | 90·2% |
| | Young_sub | 0·879 | 47·9% | 49·3% | 75·0% | 90·2% |
| 2: mid | Old | 0·836 | 21·3% | 26·6% | 59·9% | 93·6% |
| | Young | 0·813 | 10·2% | 16·5% | 53·2% | 93·6% |
| | Young_sub | 0·806 | 9·96% | 15·4% | 49·3% | 91·6% |
| 3: high | Old | 0·754 | 6·00% | 7·05% | 20·3% | 78·2% |
| | Young | 0·722 | 0·76% | 1·11% | 6·03% | 67·0% |
| | Young_sub | 0·696 | 0·77% | 1·09% | 4·49% | 40·1% |

Compared to Table S4, the percentages of true strains reaching the same cutoff values by inference always decrease in Table S5. The decrease is most pronounced in Young, shifting from being the most informative in sampling to having significantly lower coverage than Old in inference. Although the young hosts indeed include more strains, the comparison here strengthens the viewpoint that the high levels of mixed infections in these hosts hinder accurate strain inference. Conversely, inferring strains from the old hosts can provide more information on population-level strains.

### Relationship between strain abundance and recovery score

Figure S1 here is a more detailed version of Figure 3(b) in the main text. Here, we present a separate plot for each age group under each scenario, including the abundance in the subpopulations of sampled hosts as well. Generally, the abundant strains in the whole population are also relatively abundant in the sampled hosts. The recovery scores of strains with intermediate to high population-level abundance are overall higher in the hosts sampled from Old, except in the scenario with the lowest transmission intensity, where all age groups achieve similar recovery scores.

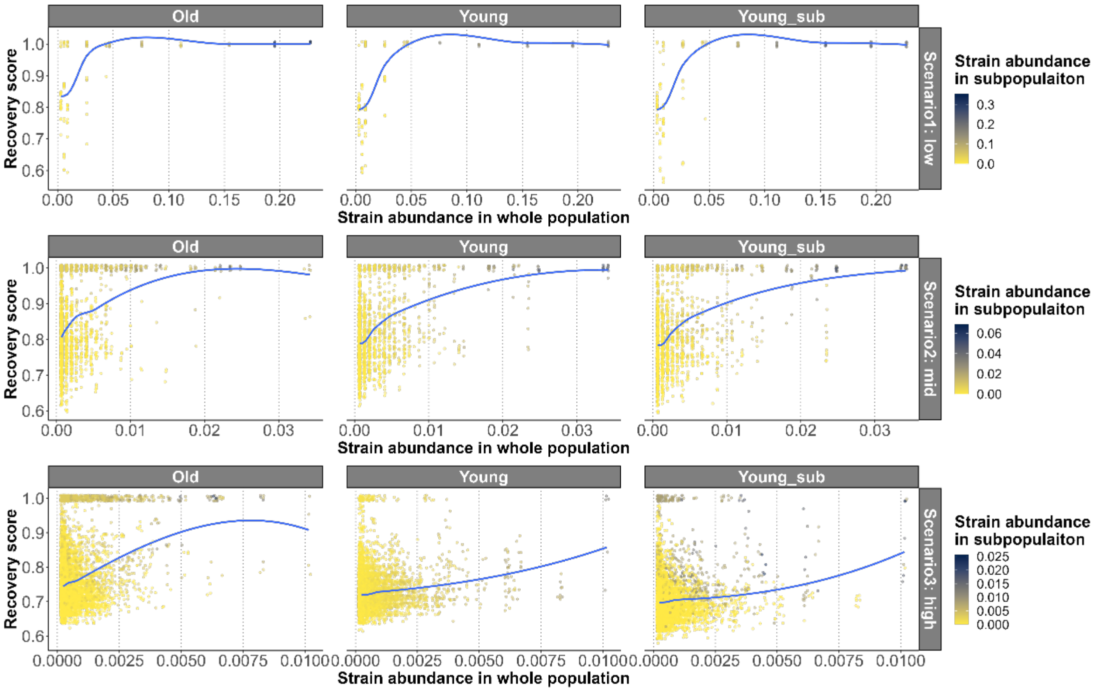

**Figure S1:** Details of the abundance of each true strain versus recovery score from inference in host subpopulations sampled from age groups Old, Young and Young_sub, corresponding to Figure 3(b) in the main text.

We also calculate an overall recovery score of all population-level true strains in each age group under each transmission intensity, as shown in Table S6. The value can be obtained by summing the products of population-level abundance and recovery score for all strains. The overall score does not appear to vary significantly among age groups, primarily due to the relatively good recovery of high-abundance strains, as illustrated in Figure S1 here and Figure 3(b) in the main text. Still, Old generally gives the highest recovery score, because of the best recovery of strains under almost all abundance levels compared to the other two age groups. Results of t-tests verify that the overall recovery scores in the hosts sampled from the Old age group are significantly higher than those in Young under all scenarios (null hypothesis: overall recovery scores in Old are no higher than those in Young; $p$=2·2e-8 for scenario 1 and $p$<2·2e-16 for scenarios 2-3).

**Table S6:** The mean and standard deviation of the overall recovery score of all true strains (sum of the products of population-level abundance and recovery score for all strains), across all 128 sampled host subpopulations in each age group under each transmission intensity.

| Scenario | Age group | Mean score | Sd score |
|---|---|---|---|
| 1: low | Old | 0·985 | 0·0070 |
| | Young | 0·981 | 0·0049 |
| | Young_sub | 0·981 | 0·0049 |
| 2: mid | Old | 0·898 | 0·0059 |
| | Young | 0·875 | 0·0078 |
| | Young_sub | 0·867 | 0·0075 |
| 3: high | Old | 0·785 | 0·0037 |
| | Young | 0·731 | 0·0041 |
| | Young_sub | 0·708 | 0·0051 |

### Effects of using different numbers of loci on inference accuracy under high transmission intensity

Since malaria has a sexual recombination stage, longer haplotypes are increasingly difficult to infer as the diversity of haplotypes will increase exponentially. In the main text, we inferred SNP-haplotypes from all 45 loci. Here, we test the inference quality using fewer loci to assess whether inference accuracy improves under fewer haplotype combinations, for the high transmission scenario (#3). Specifically, we perform inference on three consecutive non-overlapping segments (loci 1–12, 13–24, and 25–36) and in one trial on only loci 1–9 (see Supplementary Information).

Decreasing the number of loci can slightly improve the inference of within-host inference quality (WHI) and the strain dictionary (Figure S2 (a) and (b)). Meanwhile, the sampled hosts can carry or recover larger fractions of population-level true strain haplotypes when fewer loci are considered (Figure S3). The number of possible allele combinations decreases when we use fewer loci to run the inference algorithm, thus reducing the complexity to achieve higher inference accuracy. However, the effect of stochasticity may increase when too few loci are considered. Thus, for the inference of SNP-haplotypes, selection of loci needs to consider the balance between inference accuracy and the effect of randomness, and selecting an intermediate number of loci is suggested.

For the inference of shared infections between host pairs, the effect of using different numbers of loci as input is different from the case of strain haplotype inference. The general trend is that the false positive rates (1-specificity) are still lowest from Old and highest from Young, regardless of the number of loci included (Figure S2 (c)). At least, using samples from the Old age group is the least likely to get a wrong inference of shared infections. However, the effect of number of loci on true positive rates (sensitivity) is difficult to conclude. When fewer loci are used in inference, the true positive rates in Old age group decrease. Merging of the haplotypes with the same combinations of fewer selected loci leads to larger effects of stochasticity, and the inferred sharing status among strains tends to be more chaotic. When including more loci, with more information on allele combinations, the haplotypes obtained from hosts with single infections are more reliable. Therefore, the host pairs that are inferred as sharing strains are more likely to share strain in reality as well, increasing the true positive rates. The effect on host samples from Young_sub is similar to that on Old, but the small host sample sizes and the unstable inclusion of hosts with shared strains result in an even wider range for the true positive rates. On the other side, in hosts from the Young age group, when fewer loci are inferred, the true positive rates increase, but simultaneously with an increase in false positive rates, indicating a more random strain sharing pattern. The SNP-Slice algorithm tends to generate relatively small strain dictionaries with frequent strain sharing to explain the observation of allelic status. When using young hosts to infer strains, the inferred strain dictionary tends to be much smaller than the true strain dictionary (Table 2 in the main text). When fewer alleles are inferred, an even more oversimplified inferred strain dictionary can be generated with overestimated strain sharing, which is the possible reason for both high true positive rates and high false positive rates. When more loci are included in inference, at least the inferred strain dictionary size becomes larger, so both true and false positive rates drop with less inferred strain sharing, although strain sharing is still overestimated. Thus, from Figure S2 (c), the host pairwise strain-sharing network is best inferred from Old when including all 45 loci. Decreasing the number of loci may lead to less accurate or more chaotic inference. Based on these findings, we suggest including as many loci as possible to infer features on host-relatedness, such as the host pairwise strain-sharing network.

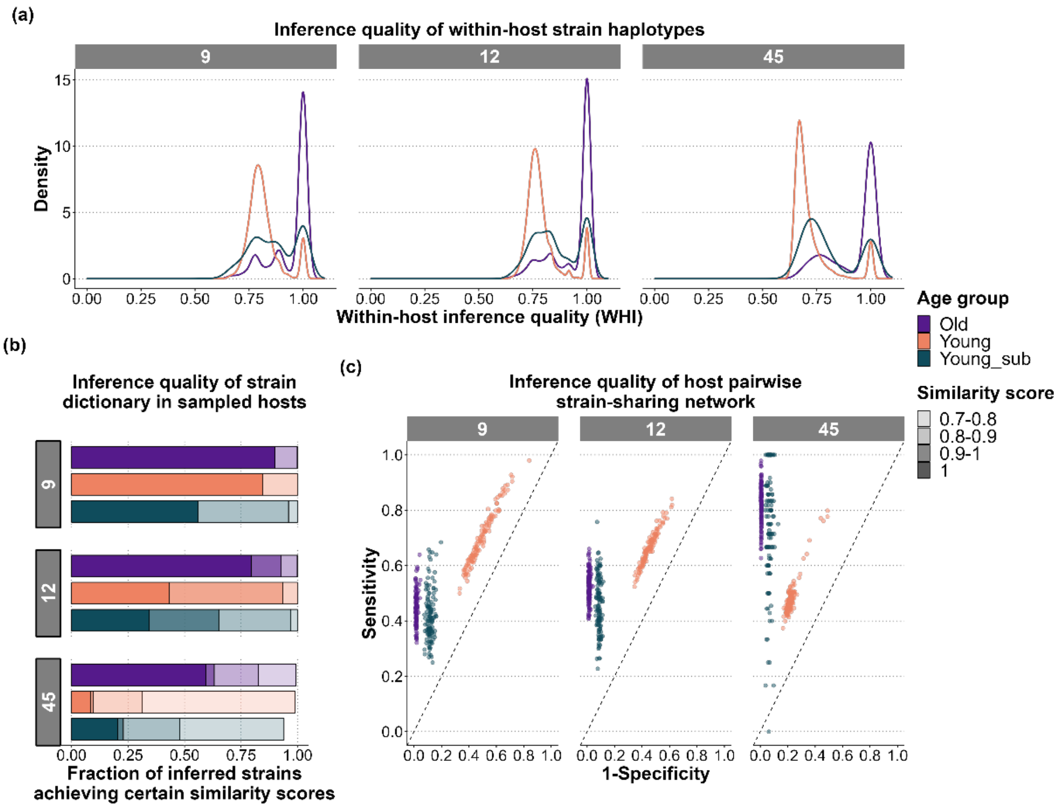

**Figure S2:** Effects of the number of loci in the inference of strain haplotypes and sharing among hosts under high transmission intensity (Scenario 3). Strain haplotypes are defined according to the combination of a certain number of loci. The plots and legends mainly follow their counterparts in Figure 2 in the main text. Here we list three loci segment lengths we input for SNP-Slice inference: 9 loci (1-9), 12 loci (combined results of inferring loci 1-12, 13-24 and 25-36), and all 45 loci. (a) The within-host inference quality, measured WHI using equation (1) in the main text. (b) The inference accuracy of the strain dictionary, shown by fractions of inferred strains achieving certain similarity scores, compared to their best matched true strains in the corresponding sampled host subsets. (c) The inference accuracy of the strain-sharing network among hosts, shown by plotting the true positive rate (sensitivity) against the false positive rate (1 – specificity).

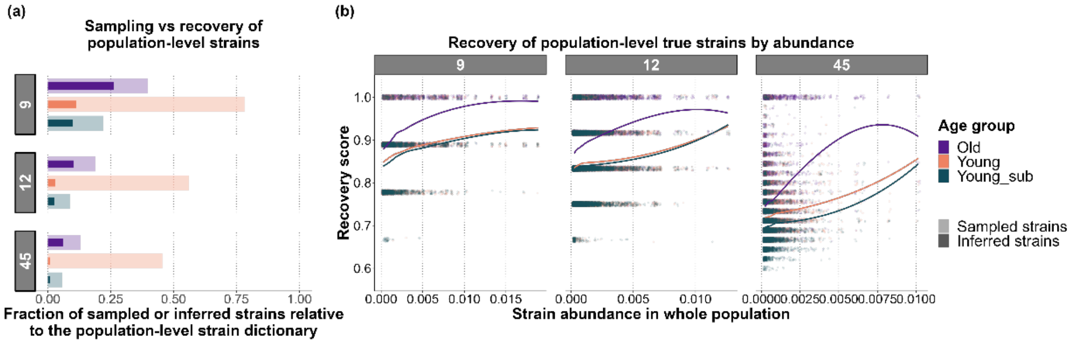

**Figure S3:** Effects of the number of loci in recovery of population-level true strains under high transmission intensity, corresponding to Figure S2. The plots and legends mainly follow their counterparts in Figure 3 in the main text. (a) Sampling and inference recovery of population-level strains in the host samples. (b) Relationship between strain abundance and recovery score. For better visualization, only 5% of the points in each case are shown in each scenario.
